# The association between major trauma centre care and outcomes of adult patients injured by low falls in England and Wales

**DOI:** 10.1101/2022.02.25.22270903

**Authors:** Michael Tonkins, Omar Bouamra, Fiona Lecky

## Abstract

**Background:** Disability and death due to low falls is increasing worldwide and disproportionately affects older adults. Current trauma systems were not designed to suit the needs of these patients. This study assessed the effectiveness of major trauma centre care in adult patients injured by low falls.

**Methods:** Data were obtained from the Trauma Audit and Research Network on adult patients injured by falls from <2 metres between 2017-2019 in England and Wales. 30-day survival, length of hospital stay and discharge destination were compared between major trauma centres (MTCs) and trauma units or local emergency hospitals (TU/LEHs).

**Results:** 127,334 patients were included of whom 27.6% attended an MTC. The median age was 79.4 years (IQR 64.5-87.2 years), and 74.2% of patients were aged >65 years. MTC care was not associated with improved 30-day survival (adjusted odds ratio [AOR] 0.91, 95% CI 0.87-0.96). Transferred patients had a significant impact upon the results. After excluding transferred patients, the AOR for survival in MTCs was 1.056 (95% CI 1.001-1.113).

**Conclusion:** TU/LEH care is at least as effective as MTC care due to the facility for secondary transfer from TU/LEHs to MTCs. In patients who are not transferred, MTCs are associated with greater odds of 30-day survival in the whole cohort and in the most severely injured patients. Future research must determine the optimum means of identifying patients in need of higher-level care; the components of care which improve patient outcomes; and develop patient-focused outcomes which reflect the characteristics and priorities of contemporary trauma patients.

**KEY MESSAGES:** *What is already known on this topic:*

- Current trauma systems were not designed to manage rising numbers of elderly patients injured by low falls.
- Previous evidence for the role of major trauma centre (MTC) care in such patients yielded conflicting results.

*What this study adds:*

- This study demonstrates that non-trauma centre care is no worse than MTC care, as long as the possibility of transfer exists.
- Therefore MTCs do have a role in the management of elderly adults injured by low falls, particularly the severely injured.

*How this study might affect research, policy or practice:*

- Research must identify those patients who need transfer, the most effective components of care, and patient-centric outcomes.

## INTRODUCTION

Trauma networks were established across England and Wales in 2012 using a ‘hub and spoke’ model in which peripheral Trauma Units (TUs) and Local Emergency Hospitals (LEHs) worked with Major Trauma Centres (MTCs) to provide trauma care to a geographically defined population.^1^ The inception of trauma networks was associated with a 19% increase in the adjusted odds of survival among severely injured patients.^1^

However, between 2000 and 2019 the mean age of a major trauma patient in England and Wales rose from 38.6 years to 61.7 years, the predominant mechanism of injury became falls from less than 2 metres (52.6%) and the prevalence of comorbidities and frailty rose.^2^ These changes represent an emerging cohort of elderly patients who sustain severe injuries in falls from standing height. Similar findings have been reported in other economically developed nations.^3–6^ There is evidence that the emerging cohort are at greater risk of being under-triaged,^7,8^ are less likely to receive care at a higher-level trauma facility,^9,10^ are more likely to be treated by junior clinicians, more likely to have longer waits for CT scans, and are less likely to be transferred to a neurosciences unit. ^11,12^

A recent systematic review has demonstrated that higher-level trauma centres are associated with lower mortality than lower-level trauma centres in patients with major trauma (adjusted odds ratio, AOR, 0.77; 95% CI 0.69-0.87).^13^ However it is unknown whether higher-level trauma centres confer the same benefit in the emerging cohort of trauma patients. The three studies that have been published on this topic were all conducted in the USA and reported divergent results.^14–16^

Therefore, the aim of this study was to establish whether there is an association between MTC care and patient outcomes after falls from less than 2 metres compared to care in a TU or LEH.

## METHODS

### Design and Setting

Data from the Trauma Audit and Research Network (TARN) covering England and Wales were analysed from 1 January 2017 - 31 December 2019. The dates were chosen to allow time for trauma network maturation and provide recent complete data.

### Population

Adult patients (aged >16 years) were eligible for inclusion if (a) they met standard TARN inclusion criteria and (b) were injured by a fall from <2 metres.

Standard TARN inclusion criteria are: admission for 3 or more nights, admission to critical care, in-hospital and emergency department deaths following trauma, and transfer to another hospital for specialist care. Important TARN exclusion criteria are: transfer for rehabilitation purposes only, isolated neck of femur or trochanteric fractures in patients aged over 65 years, and isolated closed limb fractures (except femoral fractures).

Falls from <2 metres is the TARN method of operationalising the concept of a ‘low fall’, including falls from standing height. Throughout this study reference is made to ‘older patients’: the most common threshold for ‘older’ is 65 years, and therefore this will be employed in the present study.^17,18^

### Intervention and Comparison

The intervention was care at a major trauma centre, compared to TU/LEH. Patients were classified as receiving MTC vs TU/LEH care according to the status of the first hospital they attended. Therefore patients who attended a TU with subsequent transfer to an MTC were considered in the TU cohort, and patients who attended an MTC with subsequent transfer to a TU were be considered in the MTC cohort. This is consistent with previous studies.^14–16^ The authors were not blinded to the patient group.

### Outcomes

The primary outcome was 30-day survival. 30-day survival is collected by local TARN data coordinators until hospital discharge. Deaths which occur after discharge but within 30 days of injury (the ‘true’ 30-day survival) are captured using data from the Office of National Statistics.^19^

The secondary outcomes were length of hospital stay and discharge destination. Both outcomes are recorded by local TARN data analysts at time of discharge. Length of hospital stay was measured in whole days. Discharge to the patient’s own home was compared to discharge to a location for formal or informal care (nursing home, social care, rehabilitation, other institution, home of a relative).

### Data Collection

TARN-eligible patients are identified at hospital level by local data coordinators. Patient data are prospectively captured by local coordinators using an internet-based system then checked for accuracy and completeness by trained analysts before entering the TARN database.

### Statistical Analysis

The null hypothesis for this study was that there was no association between study outcomes and MTC vs TU/LEH care.

The primary outcome of 30-day survival was analysed using binary multiple logistic regression. The regression model used the variables of TARN’s previously validated Probability of Survival (Ps) model for trauma severity: age, gender, and age x gender interaction, the modified Charleson Comorbidity Index (mCCI), the Glasgow Coma Scale (GCS), and the Injury Severity Score (ISS).^19^ Three other variables known to be associated with mortality in older trauma patients were also included in the model.^17,20^ These were the most injured body area, anticoagulation, and hypotension (systolic blood pressure <110mmHg).

The ISS is the sum of the squares of the three most severely injured body regions using the Abbreviated Injury Scale (AIS).^21^ Because the ISS is non-linear, fractional polynomial transformation was undertaken to avoid violating the assumptions of logistic regression.^19,22^

Hospital length of stay was compared in patients who were discharged to a known location. Patients who died, were transferred to another acute hospital, were discharged to no fixed abode or for whom the discharge destination was unknown were excluded. Length of stay was curtailed at 365 days to reduce the influence of outliers. Cox regression was used to adjust for the casemix between MTCs and TU/LEHs using the same predictor variables employed in the primary logistic regression model, because these are also known to be associated with hospital length of stay.^23–28^ The assumption of proportional hazards was checked by visual inspection of the survival curves.

Discharge destination was dichotomised into ‘own home’ versus any form of formal or informal care. Patients who died, remained in hospital, whose discharge destination is unknown or of no fixed abode, and those who were transferred to another facility were excluded. Binary multiple logistic regression was used to establish whether trauma centre status is associated with discharge destination. Based on previous evidence, the same predictor variables employed in the primary outcome logistic regression model were used in the discharge destination model.^26,29,30^

Missing data were imputed using multiple imputation under the assumption of missingness at random and applying Rubin’s rule on the 10 imputed sets.

### Sensitivity and Subgroup Analyses

Transfer from lower-to higher-level care has been observed to confer a survival benefit compared to remaining in lower-level care.^31,32^ Therefore a sensitivity analysis restricted to all patients who were *not* transferred between hospitals was performed in order to investigate the impact of inter-hospital transfer within trauma networks.

Most patients hospitalised due to injuries caused by low falls are aged 65 years and above, and these patients are most representative of the ‘emerging cohort’ of trauma patients discussed above. Therefore, a subgroup analysis restricted to all patients aged 65 years and above was performed.

An ISS of >15 is the most common threshold for ‘major trauma’. Strong evidence exists that higher-level care within a trauma system is associated with improved outcomes in patients with an ISS >15.^13^ Therefore this subgroup analysis was performed to establish whether MTC care continues to afford a survival advantage in the most severely injured patients who fell from <2m.

### Statistical Software

Data analysis was conducted using SPSS (version 26, IBM Corporation; Armonk, New York).

### Patient and Public Involvement

Two recent consensus-building exercises involving patients and the public report that identifying which patients benefit from major trauma centre care is an important emergency medicine research priority.^33,34^ This study was authorised by the TARN research committee which reports to the TARN Board where there is standing PPI presence.

### Ethical Approval

TARN has UK Health Research Authority Confidentiality Advisory Group approval for research using patient data under Section 251 of the NHS Act (2006). Ethical approval for the use of anonymised clinical data were approved by the Research Ethics Committee of the University of Sheffield (application reference number 037465).

## RESULTS

### Data Completeness

Data were 98.9% complete. Missing values were distributed across 10 variables. With the exception of the Glasgow Outcome Score, the difference in missingness between MTCs and TU/LEHs was <1%. Missingness of outcome data was 1.5% for 30-day survival and 0.3% for discharge destination. Length of stay was complete for all patients.

### Demographic Data and Unadjusted Outcomes

127,334 patients were included of whom 35,175 (27.6%) attended a MTC (n=29) and 92,159 (72.4%) attended a TU (n=140) or LEH (n=26) - see Figure 1. The median age was 79.4 years (IQR 64.5-87.2 years), and 74.2% of patients were aged >65 years. Most patients were female (59.1%).

**Figure 1:**
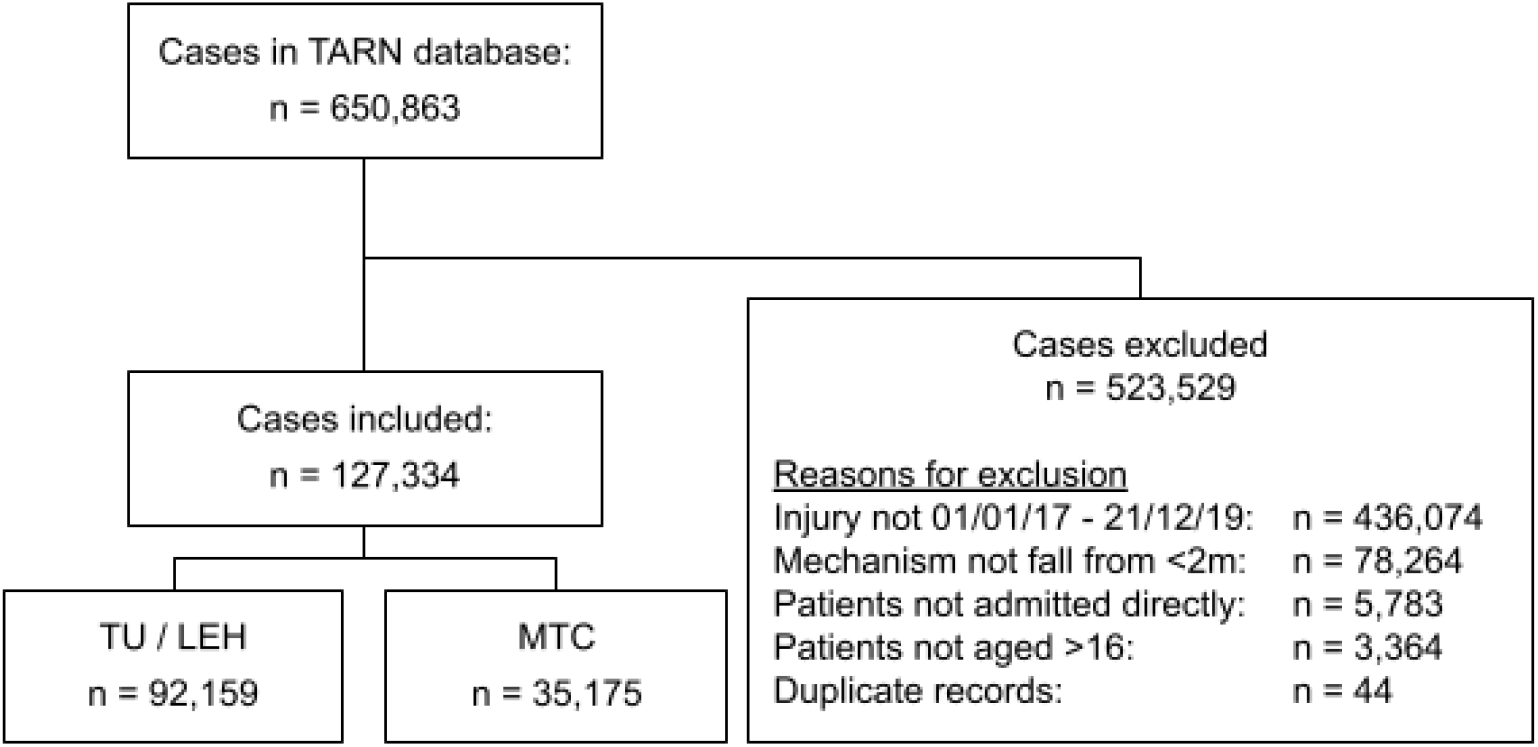
STROBE diagram.

**Figure 2:**
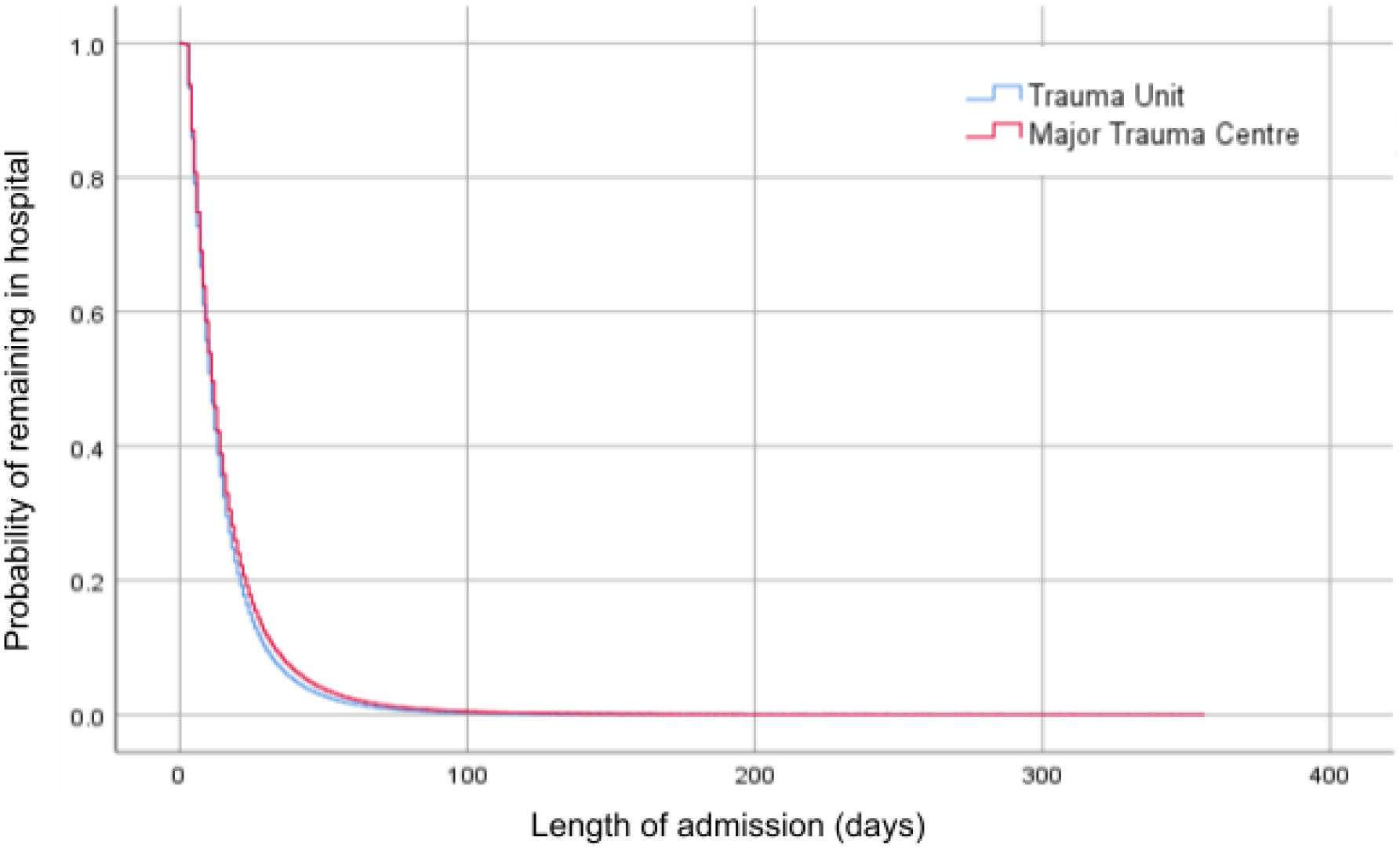
Kaplan-Meier curves for length of hospital admission, after controlling for case mix.

Between-group differences are presented in Table 1. MTC patients tended to be slightly younger, were more likely to be male, more severely injured, and suffer a higher proportion of head injuries. MTC patients were more likely to trigger trauma team activation and to undergo CT scanning, surgery, to be admitted to critical care and to receive a rehabilitation prescription.

**Table 1:** Cohort characteristics by trauma centre level.

|  |  | Trauma Unit or LEH |  | Major Trauma Centre |  | Comparison |  |
| --- | --- | --- | --- | --- | --- | --- | --- |
|  |  | 92,159 (72.4%) |  | 35,175 (27.6%) |  |  |  |
|  |  | N | % | N | % | Difference* | Significance |
| Age (years). Median, IQR |  | 79.8 | 65.0 - 87.4 | 78.3 | 63.3 - 86.7 | +1.5 years | <0.001<sup>‡</sup> |
| Female |  | 55423 | 60.1% | 19866 | 56.5% | +3.6% | <0.001 |
| mCCI band | mCCI 0 | 26729 | 29.6% | 9998 | 28.8% | +0.8% | <0.001<sup>‡</sup> |
|  | mCCI 1-5 | 41809 | 46.3% | 15502 | 44.6% | +1.7% |  |
|  | mCCI 6-10 | 17289 | 19.1% | 7054 | 20.3% | -1.2% |  |
|  | mCCI >10 | 4554 | 5.0% | 2185 | 6.3% | -1.3% |  |
| Prescribed anticoagulation |  | 12443 | 13.5% | 4685 | 13.3% | +0.2% | 0.398 |
| Patients aged 65 and over |  | 69120 | 75.0% | 25415 | 72.3% | +2.7% |  |
| ISS band | ISS 1-8 | 22445 | 24.4% | 7010 | 19.9% | +4.5% | <0.001<sup>‡</sup> |
|  | ISS 9-15 | 44930 | 48.8% | 15975 | 45.4% | +3.4% |  |
|  | ISS >15 | 24784 | 26.9% | 12190 | 34.7% | -7.8% |  |
| GCS band | GCS 3 | 466 | 0.5% | 669 | 2.0% | -1.5% | <0.001<sup>‡</sup> |
|  | GCS 4-5 | 240 | 0.3% | 304 | 0.9% | -0.6% |  |
|  | GCS 6-8 | 603 | 0.7% | 578 | 1.7% | -1.0% |  |
|  | GCS 9-12 | 1964 | 2.3% | 1372 | 4.1% | -1.8% |  |
|  | GCS 13-14 | 11804 | 13.5% | 5169 | 15.4% | -1.9% |  |
|  | GCS 15 | 72117 | 82.7% | 25486 | 75.9% | +6.8% |  |
| Most severely injured body region | Abdomen | 554 | 0.6% | 228 | 0.6% | 0.0% | <0.001 |
|  | Chest | 13402 | 14.5% | 4877 | 13.9% | +0.6% |  |
|  | Face | 576 | 0.6% | 277 | 0.8% | -0.2% |  |
|  | Head | 21719 | 23.6% | 9629 | 27.4% | -3.8% |  |
|  | Limbs | 37288 | 40.5% | 13593 | 38.6% | +1.9% |  |
|  | Multiple | 5257 | 5.7% | 2153 | 6.1% | -0.4% |  |
|  | Other | 54 | 0.1% | 43 | 0.1% | 0.0% |  |
|  | Spine | 13309 | 14.4% | 4375 | 12.4% | +2.0% |  |
| Patients with head injuries |  | 24855 | 27.0% | 11571 | 32.9% | -5.9% | <0.001 |
| Shock (SBP <110mmHg) |  | 12447 | 13.5% | 5367 | 15.3% | -1.8% | <0.001 |
| Had trauma team |  | 3745 | 4.1% | 6302 | 17.9% | -13.8% | <0.001 |
| Had CT scan |  | 60811 | 66.0% | 25647 | 72.9% | -6.9% | <0.001 |
| Had surgery |  | 25964 | 28.2% | 13036 | 37.1% | -8.9% | <0.001 |
| Had interventional radiology |  | 50 | 0.1% | 106 | 0.3% | -0.2% | <0.001 |
| Intubated and ventilated |  | 1093 | 1.2% | 1787 | 5.2% | -4.0% | <0.001 |
| Admitted to critical care |  | 3749 | 4.1% | 4155 | 12.0% | -7.9% | <0.001 |
| Patient underwent transfer |  | 8344 | 9.1% | 1184 | 3.4% | +5.7% | <0.001 |
| Rehabilitation prescription |  | 15474 | 23.3% | 23229 | 85.0% | -61.7% | <0.001 |
| Survived to 30 days |  | 85660 | 92.9% | 31708 | 90.1% | +2.8% | <0.001 |
| Length of stay in hospital (days) |  | 10 | 5 – 19 | 11 | 6 - 21 | -1 | <0.001<sup>‡</sup> |
| Length of stay in critical care (days) |  | 2 | 1 – 5 | 3 | 1 - 9 | -1 | <0.001<sup>‡</sup> |
| Discharged to own home |  | 52635 | 69.10% | 21012 | 70.30% | -1.2% | <0.001 |

Hypothesis tests are Chi-squared with Yates' Continuity Correction unless otherwise specified. \*TU-MTC †Mann-Whitney U Test.

In the unadjusted analysis of non-imputed data the odds ratio (OR) for survival in an MTC were 0.69 (95% confidence interval [CI] 0.66-0.73, χ2 p<0.001) compared to TU care. In MTCs, patients spent a median duration of 1 day more in hospital, and 1.2% more patients were discharged to their own home.

### 30-day Survival

In the logistic regression model the Adjusted Odds Ratio (AOR) of 30-day survival in MTCs compared with TU/LEHs were 0.91 (95% CI 0.87-0.96, p <0.001). This can be interpreted as a 9% decreased odds of 30-day survival in MTCs compared to TU/LEHs. The C-statistic (Area Under the Receiving Operator Characteristic Curve, AUROC) was 0.83 (95% CI 0.82-0.83, p<0.001), indicating a strong model. The full model and ROC curve are available in the supplementary material.

Patients aged over 65 years comprised 74.2% (n=32823) of the whole cohort. These patients had higher rates of comorbidity and anticoagulation. Full details are available in Supplementary Table 7. The unadjusted odds ratio for 30-day survival in patients aged >65 years who received MTC care was 0.715 (95% CI 0.683-0.749, p<0.001). The adjusted odds ratio of survival in patients aged >65 years who received MTC care was 0.929 (95% CI 0.880-0.980, p=0.007), AUROC 0.794 (95% CI 0.789-0.799).

Patients with an ISS >15 comprised 29.0% (n=36974) of the whole cohort. Their 30-day mortality was higher at 15.6% compared to 7.8% in the whole cohort. Full details are available in Supplementary Table 7. The unadjusted odds ratio of 30-day survival in patients with ISS >15 who received MTC care was 0.702 (95% CI 0.663-0.744, p<0.001). The adjusted odds ratio of survival in patients with an ISS >15 who received MTC care was 0.852 (95% CI 0.974-0.915, p<0.001), AUROC 0.821 (95% CI 0.815-0.827).

Patients who did not undergo transfer comprised 92.5% (n=117806) of the whole cohort. Compared to transferred patients, their rate of 30-day survival was lower (91.6% compared to 99.4%). Full details are available in Supplementary Table 8. Most transfers (69.8%) were TU-to-MTC, among whom the rate of 30-day survival was 99.2%. In the sensitivity analysis excluding transferred patients, the AOR of 30-day survival in MTCs compared with TU/LEHs were 1.056 (95% CI 1.001-1.113, p 0.044). Model performance (AUROC) was 0.83 (95% CI 0.829-0.838). This represents a statistically significant reversal of the primary analysis.

Given the impact of transferred patients upon the primary outcome, a post-hoc analysis was conducted in which transferred patients were excluded from the >65 years and ISS >15 subgroups. In both subgroups the result changed. Once transfers were excluded, MTC care was no longer associated with a statistically significant difference in 30-day survival in patients aged >65 years. Conversely, in patients with an ISS >15 MTC care was found to be associated with improved odds of 30-day survival (AOR 1.126, 95% CI 1.033-1.215, p 0.002). Table 2 summarises the results of all analyses and the impact of transfers upon them.

**Table 2:**
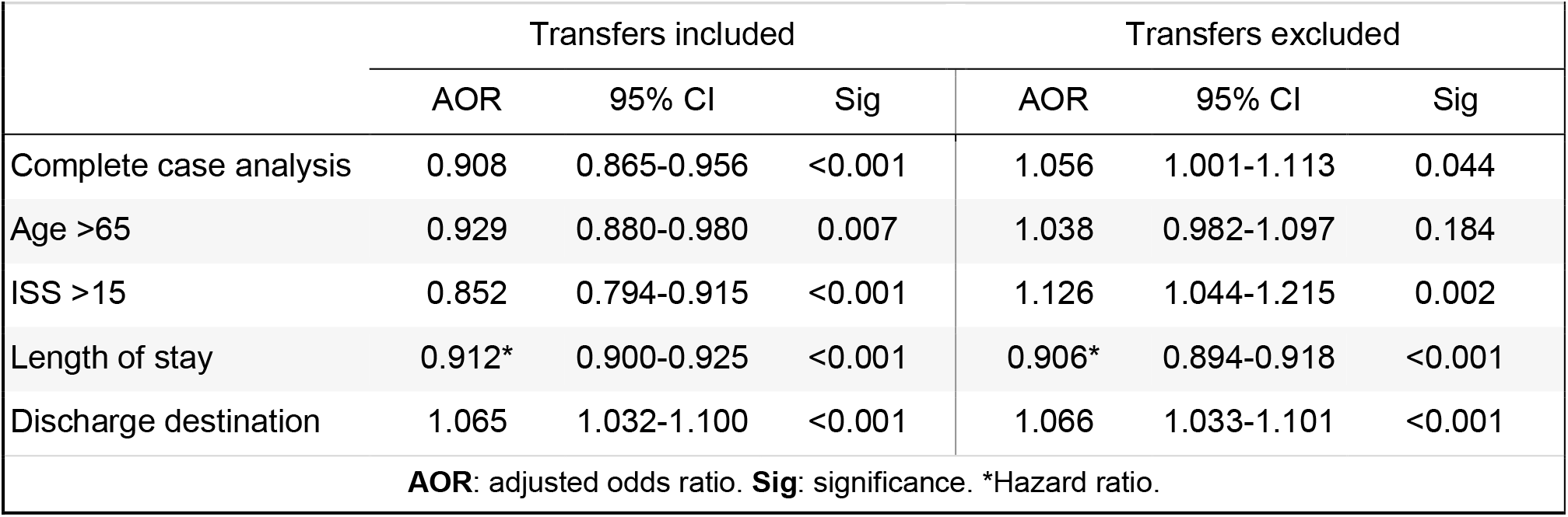
Impact of excluding transfers on the primary and secondary outcomes.

|  |  |  |  |  |  |  |
| --- | --- | --- | --- | --- | --- | --- |
|  | Transfers included |  |  | Transfers excluded |  |  |
|  | AOR | 95% CI | Sig | AOR | 95% CI | Sig |
| Complete case analysis | 0.908 | 0.865-0.956 | <0.001 | 1.056 | 1.001-1.113 | 0.044 |
| Age >65 | 0.929 | 0.880-0.980 | 0.007 | 1.038 | 0.982-1.097 | 0.184 |
| ISS >15 | 0.852 | 0.794-0.915 | <0.001 | 1.126 | 1.044-1.215 | 0.002 |
| Length of stay | 0.912* | 0.900-0.925 | <0.001 | 0.906* | 0.894-0.918 | <0.001 |
| Discharge destination | 1.065 | 1.032-1.100 | <0.001 | 1.066 | 1.033-1.101 | <0.001 |
| **AOR:** adjusted odds ratio. **Sig:** significance. *Hazard ratio. |  |  |  |  |  |  |

### Length of Stay

Overall, 83.3% of patients (n=106,014) were discharged alive to a known location within 365 days. In unadjusted analyses the median time to discharge in TUs was 11 days (IQR 6-19 days) and in MTCs was 12 days (IQR 6-21 days). In the adjusted analysis the hazard ratio for MTC care was 0.912 (95% CI 0.900-0.925, p <0.001). Exclusion of transferred patients did not produce a statistically significant result.

### Discharge Destination

Overall, 83.3% of patients were discharged either home or to an informal or formal care location. The unadjusted odds ratio for discharge home from an MTC was 1.06 (95% CI 1.029-1.091, p <0.001). After adjusting for casemix, the adjusted odds ratio of patient discharge to their own home from an MTC was 1.065 (95% CI 1.032-1.100, p <0.001). The AUROC was 0.744 (95% CI 0.739 - 0.745). Exclusion of transferred patients did not produce a statistically significant result.

## DISCUSSION

### Summary of Results

This study demonstrates lower adjusted odds of 30-day survival in adult patients injured by low falls who attended MTCs, compared to those who attended TU/LEHs (AOR 0.908, 95% CI 0.865-0.956) as the first hospital. This result persisted in patients who were over 65 years of age (AOR 0.715, 95% CI 0.683-0.749, p<0.001), and patients who had an ISS >15 (AOR 0.852, 95% CI 0.974-0.915, p<0.001).

Once transferred patients were excluded, the adjusted odds of 30-day survival were higher in MTCs than TU/LEHs (AOR 1.056, 95% CI 1.001-1.113, p 0.044). Excluding transferred patients also affected the results of patients aged >65 years, in whom MTC care was no longer associated with a statistically significant difference in odds of 30-day survival (AOR 1.038, 95% CI 0.982 - 1.097, p=0.184). The result for patients with ISS >15 also changed, and MTC care was found to be associated with higher odds of 30-day survival (AOR 1.126, 95% CI 1.044-1.215).

### Interpretation of Results

The primary analysis demonstrates that TU/LEH care is at least as effective as MTC care. However, patients who were transferred between centres had a significant impact upon the results. Most transfers (69.8%) were TU/LEH-to-MTC, and the very high rate of 30-day survival (99.2%). In the primary analysis these patients were mostly attributed to TU/LEH care, thereby censoring the input of care from further hospitals.

Excluding transferred patients reveals a more precise analysis of the association between level of care and patient outcomes. Doing so demonstrates that higher-level care is associated with improved odds of survival in the whole cohort and in the most severely injured, but not in those over 65 years old. The high rate of survival among transferred patients illustrates that initial attendance at an MTC is not necessary, providing that the option to access MTC care via secondary transfer remains. This finding demonstrates the importance of early injury identification, collaboration between care providers and capacity planning within trauma systems.

### Relation to Previous Research

The results of this study are concordant with the recent systematic review by Van Ditshuizen et al.^13^ Firstly, in the primary analysis higher-level (MTC) care was not found to be associated with a survival benefit in the general population. Secondly, failure to exclude transferred patients results in underestimation of the survival benefit at higher-level care. Thirdly, higher-level care is associated with a survival benefit for patients with major trauma (ISS >15).

The lack of a statistically significant association between MTC care and 30-day survival in patients aged >65 years suggests that the benefits of MTC care become attenuated with increasing age. This is concordant with the results of Ahmed et al^16^, who found no association between higher-level care and survival in patients aged >65 years injured by ground-level falls. It is probable that the accumulated mortality risk attributable to age, comorbidities and frailty is a major cause for this finding. However, in the face of widespread evidence that older patients receive suboptimal care at multiple points in their pathway, it is likely that there are opportunities to improve outcomes in this patient group.

### Implications for Future Research

The results of this study highlight three areas for further research. Firstly, tools to identify which patients benefit from MTC care must be developed and validated with associated decision-analytic modelling. Secondly, the components and characteristics of MTC care associated with a survival benefit should be identified, developed, and where possible distributed throughout trauma networks. Thirdly, specific outcome measures for older trauma patients must be developed.

### Strengths and Weaknesses

This is the first study in England and Wales to examine outcomes of patients injured in falls from standing height cared for at MTCs compared to TU/LEHs. The result of this study provides evidence for the evaluation of the role of MTC care for these patients.

The results of this study remain at risk of selection bias and confounding due to unmeasured baseline factors including patient frailty. However, as there was no clinically significant difference in age or comorbidity (both strongly associated with frailty measures) between the comparator groups this may not be a serious flaw. No statistical analysis plan was made publicly available in advance beyond the TARN Research Committee, impacting the transparency of the study and increasing risk of reporting bias. The investigators were not blinded to the treatment group. However, the data were supplied under a data sharing agreement from TARN for the study objectives. Beyond imputation for missing variables the dataset was not amended.

## CONCLUSION

By focusing upon adult patients who fall from standing height, this study has sought to capture the overlapping characteristics of mechanism, age and comorbidities which defines the emerging cohort of trauma patients and thereby to determine, in part, the role of MTCs in their care. In the primary analysis, TU/LEH care was found to be at least as effective as MTC care due to the facility within trauma networks for secondary transfer from lower-level to higher-level care. In patients who were not transferred, MTCs were found to be associated with greater odds of 30-day survival in the whole cohort and in the most severely injured patients, but here was no association between MTC care and improved 30-day survival in patients aged >65 years.

## Data Availability

All data produced in the present study are available upon reasonable request to the authors.

## Acknowledgements

The authors would like to acknowledge the essential work by Shiraz Ahmed and Laura White of TARN in preparing and supplying the data.

