## Supplementary for "The association between major trauma centre care and outcomes of adult patients injured by low falls in England and Wales"

**Tables In Main Body of Manuscript**

Figure 1 - STROBE diagram
Table 1 - Baseline cohort characteristics.
Table 2 - Summary of mortality results
Figure 2 - Kaplan-Meier curves for LOS

**Supplementary Tables**

Missing Data

Supplementary Table 1- Incomplete Variables
Supplementary Table 2 - Incomplete & Complete Cases
Supplementary Table 3 - Incompleteness by TC status
Supplementary Table 4 - Original and imputed values

Primary Analysis

Supplementary Table 5 - Variables used in logistic regression modelling
Supplementary Table 6 – Primary analysis multiple logistic regression model
Supplementary Figure 1- ROC Curve for primary logistic regression model
Supplementary Table 7 - Characteristics of subgroup analysis cohorts
Supplementary Table 8 - Characteristics of transferred and non-transferred patients
Supplementary Table 9 - Characteristics of transferred patients by direction of transfer

Secondary Analysis

Supplementary Table 10 - Discharge destination by trauma centre status
Supplementary Table 11 - Logistic regression model for discharge destination
Supplementary Table 12 - Cox regression model for length of stay

### Tables

| Supplementary Table 1: Variables with missing data points | | | |
|---|---|---|---|
| | Missing values within variable | Percentage of variable missing | Percentage of all missing data |
| Glasgow Outcome Score | 24897 | 19.6% | 61.8% |
| Glasgow Coma Scale | 6562 | 5.2% | 16.3% |
| Modified Charleson Comorbidity Index | 2214 | 1.7% | 5.5% |
| Intubated and ventilated | 1879 | 1.5% | 4.7% |
| 30 day survival | 1873 | 1.5% | 4.6% |
| Length of stay on critical care | 1658 | 1.3% | 4.1% |
| Time to CT | 677 | 0.5% | 1.7% |
| Discharge destination | 333 | 0.3% | 0.8% |
| Time to surgery | 172 | 0.1% | 0.4% |
| Rehabilitation prescription | 30 | 0.0% | 0.1% |

**Supplementary Table 2: Characteristics of incomplete and complete cases.**

| | | **Incomplete Cases** 32780 (25.7%) | | **Complete Cases** 94554 (74.3%) | | **Comparison** | | | |
|---|---|---|---|---|---|---|---|---|---|
| | | n | % | n | % | Diff | OR | 95% CI | Sig |
| Major Trauma Centre | | 6133 | 18.7% | 29042 | 30.7% | 12.0% | 0.591 | 0.503 - 0.536 | <0.000 |
| Age (years). Median (IQR). | | 77.8 | 62.8-86.4 | 79.9 | 65.3-87.4 | 2.1 | - | - | <0.000干 |
| Female | | 18845 | 57.5% | 56444 | 59.7% | 2.2% | 0.913 | 0.890 - 0.937 | <0.000 |
| mCCI bands | 0 | 9099 | 29.8% | 27628 | 29.2% | 0.6% | - | - | <0.592干 |
| | 1-5 | 13830 | 45.2% | 43481 | 46.0% | 0.8% | | | |
| | 6-10 | 5899 | 19.3% | 18444 | 19.5% | 0.2% | | | |
| | >10 | 1738 | 5.7% | 5001 | 5.3% | 0.4% | | | |
| ISS bands | 1-8 | 7776 | 23.7% | 21679 | 22.9% | 0.8% | - | - | <0.000干 |
| | 9-15 | 14874 | 45.4% | 46031 | 48.7% | 3.3% | | | |
| | >15 | 10130 | 30.9% | 26844 | 28.4% | 2.5% | | | |
| Major trauma (ISS >15) | | 10130 | 30.9% | 26844 | 28.4% | 2.5% | 0.886 | 0.863 - 0.911 | <0.000 |
| GCS bands | 3 | 181 | 0.7% | 954 | 1.0% | 0.3% | - | - | <0.185干 |
| | 4-5 | 98 | 0.4% | 446 | 0.5% | 0.1% | | | |
| | 6-8 | 231 | 0.9% | 950 | 1.0% | 0.1% | | | |
| | 9-12 | 751 | 2.9% | 2585 | 2.7% | 0.2% | | | |
| | 13-14 | 3711 | 14.2% | 13262 | 14.0% | 0.2% | | | |
| | 15 | 21246 | 81.0% | 76357 | 80.8% | 0.2% | | | |
| Shock | | 4071 | 12.4% | 13743 | 14.5% | 2.1% | 1.199 | 1.155 - 1.245 | <0.000 |
| Transferred | | 5984 | 18.3% | 3544 | 3.7% | 14.6% | 0.174 | 0.167 - 0.182 | <0.000 |
| 30-day survival | | 29874 | 96.7% | 85621 | 90.60% | 6.1% | 0.312 | 0.292 - 0.333 | <0.000 |

Hypothesis tests are Chi-squared with Yates' Continuity Correction unless otherwise specified. 干Mann-Whitney U Test.
**Diff**: difference. **OR**: odds ratio. **Sig**: significance.

| Supplementary Table 3: Data incompleteness by trauma status of first hospital | | | | | | | | |
|---|---|---|---|---|---|---|---|---|
| | **Trauma Unit / LEH** | | **Major Trauma Centre** | | **Comparison** | | | |
| | n | % | n | % | Diff | OR | 95% CI | Sig |
| GOS | 21005 | 22.8% | 3892 | 11.1% | 11.7% | 2.373 | 2.287 - 2.461 | <0.000 |
| GCS | 4965 | 5.4% | 1597 | 4.5% | 0.9% | 1.197 | 1.130 - 1.268 | <0.000 |
| mCCI | 1778 | 1.9% | 436 | 1.2% | 0.7% | 1.568 | 1.411 - 1.742 | <0.000 |
| Intubated | 1187 | 1.3% | 692 | 2.0% | 0.7% | 0.65 | 0.592 - 0.715 | <0.000 |
| Final outcome known | 1182 | 1.3% | 691 | 2.0% | 0.7% | 0.648 | 0.590 - 0.713 | <0.000 |
| Critical care length of stay | 1114 | 1.2% | 544 | 1.5% | 0.3% | 0.779 | 0.703 - 0.864 | <0.000 |
| Time to CT | 461 | 0.5% | 216 | 0.6% | 0.1% | 0.814 | 0.692 - 0.957 | <0.000 |
| Discharge destination | 292 | 0.3% | 41 | 0.1% | 0.2% | 2.724 | 1.964 - 3.778 | <0.000 |
| Time to surgery | 116 | 0.1% | 56 | 0.2% | 0.1% | 0.791 | 0.575 - 1.088 | 0.173 |
| Rehabilitation prescription | 2 | 0.0% | 28 | 0.1% | 0.1% | 0.027 | 0.006 - 0.114 | <0.000 |

All significance tests are Chi-squared with Yates' continuity correction.
**Diff**: difference. **OR**: odds ratio. **Sig**: significance.

| Supplementary Table 4: Original and imputed values | | | | | | |
|---|---|---|---|---|---|---|
| **Variable** | **Missing values (n)** | **Imputed values (n)** | **Value Distribution** | | | |
| | | | Category | Original | Imputed | Difference |
| mCCI | 1873 | 18730 | mCCI 0 | 28.8% | 29.4% | 0.6% |
| | | | mCCI 1-5 | 45.0% | 45.8% | 0.8% |
| | | | mCCI 6-10 | 19.1% | 19.4% | 0.3% |
| | | | mCCI >10 | 5.3% | 5.4% | 0.1% |
| GCS | 2214 | 22140 | GCS 3 | 0.9% | 1.1% | 0.2% |
| | | | GCS 4-5 | 0.4% | 0.5% | 0.1% |
| | | | GCS 6-8 | 0.9% | 1.0% | 0.1% |
| | | | GCS 9-12 | 2.6% | 2.8% | 0.2% |
| | | | GCS 13-14 | 13.3% | 14.0% | 0.7% |
| | | | GCS 15 | 76.7% | 80.6% | 3.9% |
| 30-day Outcome | 6562 | 65620 | Died | 7.8% | 7.9% | 0.1% |
| | | | Survived | 90.7% | 92.1% | 1.4% |

**mCCI**: Modified Charleson Comorbidity Index. **GCS**: Glasgow Coma Scale.

| Supplementary Table 5: Variables used in logistic regression modelling | | | |
|---|---|---|---|
| **Variable** | **Format** | **Values** | **Role in Imputation** |
| Age | Ordinal | 4 (16-44), 5 (45-54), 6 (55-64) 7 (65-74), 8 (75-85), 9 (>85) | Independent |
| Sex | Binary | Female, Male | Independent |
| Age*Sex | Interaction | 4, 5, 6, 7, 8, 9 | Independent |
| mCCI | Ordinal | 0, 1 (1-5), 2 (6-10), 3 (≥11) | Independent, dependent |
| Anticoagulation | Binary | Anticoagulated, not anticoagulated | Independent |
| ISS, first transformation | Scale | - | Independent |
| ISS, second transformation | Scale | - | Independent |
| GCS | Ordinal | 1 (3), 2 (4-5), 3 (6-8), 4 (9-12), 5 (13-14), 6 (15) | Independent, dependent |
| Most injured body region | Nominal | Head, abdomen, thorax, pelvis, limbs, face, spine, other | Independent |
| Head injury | Binary | Head injury, no head injury | Independent |
| Shock | Binary | Shock, no shock | Independent |
| Transfer | Binary | Transfer, no transfer | Independent |
| 30 day survival | Binary | Survived, died | Independent, dependent |
| Trauma centre status | Binary | TU, MTC | Independent |

mCCI: Modified Charleson Comorbidity Index. ISS: Injury Severity Score. Shock: Systolic Blood Pressure <110mmHg. GCS: Glasgow Coma Scale. TU: trauma unit. MTC: major trauma centre.

| Supplementary Table 6: Primary analysis multiple logistic regression model | | | | | | |
|---|---|---|---|---|---|---|
| Variable | Categories | Coefficient | Significance | AOR | 95% CI (lower) | 95% CI (upper) |
| Age | 16 - 44 (reference) | | | | | |
| | 45 - 54 | -0.341 | 0.166 | 0.711 | 0.439 | 1.153 |
| | 55 - 64 | -1.059 | 0 | 0.347 | 0.224 | 0.536 |
| | 65 - 74 | -1.672 | 0 | 0.188 | 0.123 | 0.287 |
| | 75 - 85 | -2.152 | 0 | 0.116 | 0.077 | 0.176 |
| | >85 | -2.726 | 0 | 0.065 | 0.043 | 0.099 |
| Sex | Female (reference) | | | | | |
| | Male | 0.146 | 0.55 | 1.157 | 0.717 | 1.866 |
| Age * Sex interaction | 16 - 44 * Male (reference) | | | | | |
| | 45 - 54 * Male | -0.421 | 0.155 | 0.656 | 0.368 | 1.172 |
| | 55 - 64 * Male | -0.09 | 0.733 | 0.914 | 0.543 | 1.536 |
| | 65 - 74 * Male | -0.24 | 0.349 | 0.787 | 0.476 | 1.3 |
| | 75 - 85 * Male | -0.405 | 0.101 | 0.667 | 0.411 | 1.083 |
| | >85 * Male | -0.421 | 0.089 | 0.657 | 0.405 | 1.066 |
| mCCI | 0 (reference) | | | | | |
| | 1 - 5 | -0.505 | 0 | 0.603 | 0.561 | 0.649 |
| | 6 - 10 | -0.908 | 0 | 0.403 | 0.373 | 0.436 |
| | >10 | -1.357 | 0 | 0.257 | 0.233 | 0.284 |
| Anticoagulated | Not anticoagulated (ref) | | | | | |
| | Prescribed anticoagulant | -0.095 | 0.002 | 0.91 | 0.857 | 0.966 |
| ISS | $(ISS/10)^2$ | -0.402 | 0 | 0.669 | 0.644 | 0.695 |
| | $Log(ISS/10) \times (ISS/10)^2$ | 0.194 | 0 | 1.215 | 1.183 | 1.247 |
| GCS | 15 (reference) | | | | | |
| | 3 | -3.882 | 0 | 0.021 | 0.017 | 0.025 |
| | 4 - 5 | -3.7 | 0 | 0.025 | 0.019 | 0.032 |
| | 6 - 8 | -2.686 | 0 | 0.068 | 0.059 | 0.079 |
| | 9 - 12 | -1.682 | 0 | 0.186 | 0.17 | 0.204 |
| | 13 - 14 | -0.463 | 0 | 0.629 | 0.593 | 0.667 |
| Shock | No shock (reference) | | | | | |
| | Shock | -0.649 | 0 | 0.523 | 0.493 | 0.554 |
| Injury distribution | Abdomen (reference) | | | | | |
| | Chest | -0.128 | 0.44 | 0.88 | 0.636 | 1.217 |
| | Face | 0.226 | 0.385 | 1.254 | 0.753 | 2.087 |
| | Head | 0.202 | 0.22 | 1.224 | 0.886 | 1.69 |
| | Limbs | 0.168 | 0.309 | 1.183 | 0.856 | 1.635 |
| | Multiple | 0.058 | 0.732 | 1.06 | 0.759 | 1.481 |
| | Other | -0.633 | 0.063 | 0.531 | 0.272 | 1.035 |
| | Spine | -0.031 | 0.855 | 0.97 | 0.699 | 1.345 |
| Trauma centre status | Trauma Unit (reference) | | | | | |
| | Major Trauma Centre | -0.095 | 0 | 0.91 | 0.865 | 0.957 |

| | | | | | |
|---|---|---|---|---|---|
| Constant | 6.342 | 0 | 567.746 | 337.344 | 955.511 |

**AOR**: adjusted odds ratio. **CI**: confidence interval.

**Supplementary Figure 1: Receiver operating characteristic (ROC) curve for the primary logistic regression model**

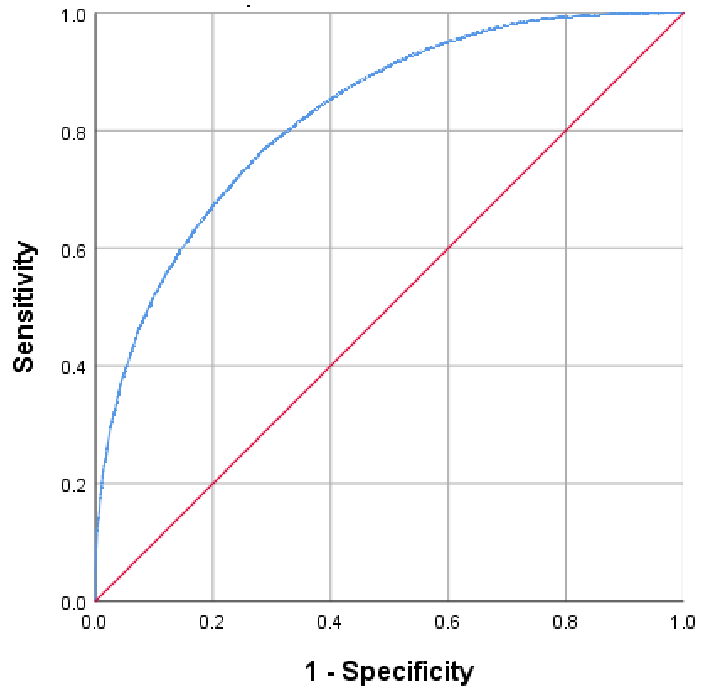

### Supplementary Table 7: Characteristics of subgroup analysis cohorts

| | | Whole Cohort 127334 (100%) | | Patients aged >65 94535 (74.2%) | | Patients with ISS >15 36974 (29.0%) | |
|---|---|---|---|---|---|---|---|
| | | n | % | n | % | n | % |
| Age (years). Median, IQR | | 79.4 | 64.5 - 87.2 | 83.7 | 76.9 - 89.0 | 80.8 | 69.1 - 87.6 |
| Female | | 75289 | 59.1% | 59249 | 62.7% | 17962 | 48.6% |
| mCCI band | mCCI 0 | 36727 | 29.4% | 22244 | 23.9% | 8377 | 23.1% |
| | mCCI 1-5 | 57311 | 45.8% | 44209 | 47.5% | 17173 | 47.3% |
| | mCCI 6-10 | 24343 | 19.5% | 21612 | 23.2% | 8322 | 22.9% |
| | mCCI >10 | 6739 | 5.4% | 5049 | 5.4% | 2457 | 6.8% |
| Prescribed anticoagulation | | 17128 | 13.5% | 15848 | 16.8% | 6339 | 17.1% |
| Patients aged 65 and over | | 94535 | 74.2% | 94535 | 100.0% | 29481 | 79.7% |
| ISS band | ISS 1-8 | 29455 | 23.1% | 24004 | 25.4% | 0 | 0.0% |
| | ISS 9-15 | 60905 | 47.8% | 41050 | 43.4% | 0 | 0.0% |
| | ISS >15 | 36974 | 29.0% | 29481 | 31.2% | 36974 | 100.0% |
| GCS band | GCS 3 | 1135 | 0.9% | 682 | 0.8% | 963 | 2.7% |
| | GCS 4-5 | 544 | 0.5% | 374 | 0.4% | 518 | 1.5% |
| | GCS 6-8 | 1181 | 1.0% | 782 | 0.9% | 1016 | 2.9% |
| | GCS 9-12 | 3336 | 2.8% | 2414 | 2.7% | 2394 | 6.8% |
| | GCS 13-14 | 16973 | 14.1% | 14514 | 16.1% | 8400 | 23.9% |
| | GCS 15 | 97603 | 80.8% | 71206 | 79.1% | 21893 | 62.2% |
| Most severely injured body region | Abdomen | 782 | 0.6% | 424 | 0.4% | 235 | 0.6% |
| | Chest | 18279 | 14.4% | 14490 | 15.3% | 4852 | 13.1% |
| | Face | 853 | 0.7% | 541 | 0.6% | 0 | 0.0% |
| | Head | 31348 | 24.6% | 25143 | 26.6% | 26813 | 72.5% |
| | Limbs | 50881 | 40.0% | 33790 | 35.7% | 2404 | 6.5% |
| | Multiple | 7410 | 5.8% | 6083 | 6.4% | 1548 | 4.2% |
| | Other | 97 | 0.1% | 64 | 0.1% | 48 | 0.1% |
| | Spine | 17684 | 13.9% | 14000 | 14.8% | 1074 | 2.9% |
| Patients with head injuries | | 36426 | 28.6% | 29384 | 31.1% | 28326 | 76.6% |
| Shock (SBP <110mmHg) | | 17814 | 14.0% | 12014 | 12.7% | 5055 | 13.7% |
| Major Trauma Centre | | 35175 | 27.6% | 25415 | 26.9% | 12190 | 33.0% |
| Had trauma team | | 10047 | 7.9% | 6442 | 6.8% | 5154 | 13.9% |
| Had CT scan | | 86458 | 67.9% | 67799 | 71.7% | 35749 | 96.7% |
| Had surgery | | 39000 | 30.6% | 23550 | 24.9% | 3916 | 10.6% |
| Had interventional radiology | | 156 | 0.1% | 105 | 0.1% | 99 | 0.3% |
| Intubated and ventilated | | 2880 | 2.3% | 1507 | 1.6% | 2492 | 6.9% |
| Admitted to critical care | | 7904 | 6.3% | 4863 | 5.2% | 4295 | 11.8% |
| Patient underwent transfer | | 9528 | 7.5% | 6130 | 6.5% | 5300 | 14.3% |
| Rehabilitation prescription | | 38703 | 41.3% | 27754 | 40.2% | 11082 | 43.8% |
| Survived to 30 days | | 117368 | 92.2% | 85423 | 90.4% | 31191 | 84.4% |
| Length of hospital stay (days) | | 10 | 5 - 19 | 12 | 6 - 21 | 9 | 4 - 19 |
| Length of critical care stay (days) | | 3 | 1 - 7 | 3 | 1 - 6 | 4 | 1 - 9 |
| Discharged to own home | | 73647 | 69.4% | 49297 | 63.3% | 17430 | 68.4% |

**Supplementary Table 8: Characteristics of transferred and non-transferred patients**

| | | Whole Cohort 127334 (100%) | | Transfers excluded 117806 (92.5%) | | Transferred patients 9528 (7.5%) | |
|---|---|---|---|---|---|---|---|
| | | n | % | n | % | n | % |
| Age (years). Median, IQR | | 79.4 | 64.5 - 87.2 | 79.9 | 65.0 - 87.5 | 72.7 | 58.4 - 82.9 |
| Female | | 75289 | 59.1% | 70873 | 60.2% | 4416 | 46.30% |
| mCCI band | mCCI 0 | 36727 | 29.4% | 33918 | 29.3% | 2809 | 29.70% |
| | mCCI 1-5 | 57311 | 45.8% | 53143 | 45.9% | 4168 | 44.10% |
| | mCCI 6-10 | 24343 | 19.5% | 22496 | 19.4% | 1847 | 19.50% |
| | mCCI >10 | 6739 | 5.4% | 6112 | 5.3% | 627 | 6.60% |
| Prescribed anticoagulation | | 17128 | 13.5% | 16088 | 13.7% | 1040 | 10.90% |
| Patients aged 65 and over | | 94535 | 74.2% | 88405 | 75.0% | 6130 | 64.30% |
| ISS band | ISS 1-8 | 29455 | 23.1% | 28303 | 24.0% | 1152 | 12.10% |
| | ISS 9-15 | 60905 | 47.8% | 57829 | 49.1% | 3076 | 32.30% |
| | ISS >15 | 36974 | 29.0% | 31674 | 26.9% | 5300 | 55.60% |
| GCS band | GCS 3 | 1135 | 0.9% | 1013 | 0.9% | 122 | 1.30% |
| | GCS 4-5 | 544 | 0.5% | 471 | 0.4% | 73 | 0.80% |
| | GCS 6-8 | 1181 | 1.0% | 1001 | 0.9% | 180 | 1.90% |
| | GCS 9-12 | 3336 | 2.8% | 2861 | 2.6% | 475 | 5.10% |
| | GCS 13-14 | 16973 | 14.1% | 15447 | 13.9% | 1526 | 16.40% |
| | GCS 15 | 97603 | 80.8% | 90654 | 81.3% | 6949 | 74.50% |
| Most severely injured body region | Abdomen | 782 | 0.6% | 663 | 0.6% | 119 | 1.20% |
| | Chest | 18279 | 14.4% | 17295 | 14.7% | 984 | 10.30% |
| | Face | 853 | 0.7% | 753 | 0.6% | 100 | 1.00% |
| | Head | 31348 | 24.6% | 27159 | 23.1% | 4189 | 44.00% |
| | Limbs | 50881 | 40.0% | 48579 | 41.2% | 2302 | 24.20% |
| | Multiple | 7410 | 5.8% | 6917 | 5.9% | 493 | 5.20% |
| | Other | 97 | 0.1% | 89 | 0.1% | 8 | 0.10% |
| | Spine | 17684 | 13.9% | 16351 | 13.9% | 1333 | 14.00% |
| Patients with head injuries | | 36426 | 28.6% | 31794 | 27.0% | 4632 | 48.60% |
| Shock (SBP <110mmHg) | | 17814 | 14.0% | 16425 | 13.9% | 1389 | 14.60% |
| Major Trauma Centre | | 35175 | 27.6% | 33991 | 28.9% | 1184 | 12.40% |
| Had trauma team | | 10047 | 7.9% | 8744 | 7.4% | 1303 | 13.70% |
| Had CT scan | | 86458 | 67.9% | 78720 | 66.8% | 7738 | 81.20% |
| Had surgery | | 39000 | 30.6% | 38130 | 32.4% | 870 | 9.10% |
| Had interventional radiology | | 156 | 0.1% | 136 | 0.1% | 20 | 0.20% |
| Intubated and ventilated | | 2880 | 2.3% | 2241 | 1.9% | 639 | 7.90% |
| Admitted to critical care | | 7904 | 6.3% | 7305 | 6.2% | 599 | 7.20% |
| Patient underwent transfer | | 9528 | 7.5% | 0 | 0.0% | 9528 | 100.00% |
| Rehabilitation prescription | | 38703 | 41.3% | 37310 | 42.1% | 1393 | 27.20% |
| Survived to 30 days | | 117368 | 92.2% | 107901 | 91.6% | 9467 | 99.40% |
| Length of hospital stay (days) | | 10 | 5 - 19 | 11 | 6 - 20 | 2 | 1 - 11 |
| Length of critical care stay (days) | | 3 | 1 - 7 | 3 | 1 - 7 | 4 | 1 - 10 |
| Discharged to own home | | 73647 | 69.4% | 73420 | 69.5% | 227 | 64.70% |

| Supplementary Table 9: Characteristics of transferred patients by direction of transfer | | | | | | | | | |
|---|---|---|---|---|---|---|---|---|---|
| | | **TU/LEH to MTC** 6653 (69.8%) | | **MTC to TU/LEH** 927 (9.7%) | | **MTC to MTC** 257 (2.7%) | | **TU/LEH to TU/LEH** 1691 (17.7%) | |
| | | n | % | n | % | n | % | n | % |
| Age (years). Median, IQR | | 71.8 | 57.4 - 81.9 | 77.5 | 62.9 - 85.7 | 59.8 | 46.0 - 73.4 | 76.2 | 62.2 - 85.0 |
| Female | | 2899 | 43.6% | 504 | 54.4% | 88 | 34.2% | 925 | 54.7% |
| mCCI band | mCCI 0 | 1966 | 29.7% | 251 | 27.6% | 84 | 32.9% | 508 | 30.3% |
| | mCCI 1-5 | 2924 | 44.2% | 408 | 44.9% | 106 | 41.6% | 730 | 43.6% |
| | mCCI 6-10 | 1259 | 19.0% | 201 | 22.1% | 49 | 19.2% | 338 | 20.2% |
| | mCCI >10 | 463 | 7.0% | 48 | 5.3% | 16 | 6.3% | 100 | 6.0% |
| Prescribed anticoagulation | | 728 | 10.9% | 103 | 11.1% | 20 | 7.8% | 189 | 11.2% |
| Patients aged 65 and over | | 4181 | 62.8% | 661 | 71.3% | 103 | 40.1% | 1185 | 70.1% |
| ISS band | ISS 1-8 | 731 | 11.0% | 69 | 7.4% | 13 | 5.1% | 339 | 20.0% |
| | ISS 9-15 | 1775 | 26.7% | 402 | 43.4% | 55 | 21.4% | 844 | 49.9% |
| | ISS >15 | 4147 | 62.3% | 456 | 49.2% | 189 | 73.5% | 508 | 30.0% |
| GCS band | GCS 3 | 91 | 1.4% | 21 | 2.4% | 5 | 2.0% | 5 | 0.3% |
| | GCS 4-5 | 53 | 0.8% | 14 | 1.6% | 4 | 1.6% | 2 | 0.1% |
| | GCS 6-8 | 126 | 1.9% | 39 | 4.4% | 11 | 4.4% | 4 | 0.2% |
| | GCS 9-12 | 335 | 5.1% | 84 | 9.4% | 22 | 8.8% | 34 | 2.1% |
| | GCS 13-14 | 1083 | 16.5% | 185 | 20.8% | 55 | 21.9% | 203 | 12.5% |
| | GCS 15 | 4874 | 74.3% | 548 | 61.5% | 154 | 61.4% | 1373 | 84.7% |
| Most severely injured body region | Abdomen | 97 | 1.5% | 4 | 0.4% | 1 | 0.4% | 17 | 1.0% |
| | Chest | 663 | 10.0% | 71 | 7.7% | 6 | 2.3% | 244 | 14.4% |
| | Face | 50 | 0.8% | 4 | 0.4% | 0 | 0.0% | 46 | 2.7% |
| | Head | 3272 | 49.2% | 334 | 36.0% | 167 | 65.0% | 416 | 24.6% |
| | Limbs | 1337 | 20.1% | 332 | 35.8% | 19 | 7.4% | 614 | 36.3% |
| | Multiple | 308 | 4.6% | 74 | 8.0% | 11 | 4.3% | 100 | 5.9% |
| | Other | 5 | 0.1% | 0 | 0.0% | 1 | 0.4% | 2 | 0.1% |
| | Spine | 921 | 13.8% | 108 | 11.7% | 52 | 20.2% | 252 | 14.9% |
| Patients with head injuries | | 3554 | 53.4% | 421 | 45.4% | 186 | 72.4% | 471 | 27.9% |
| Shock (SBP <110mmHg) | | 982 | 14.8% | 160 | 17.3% | 51 | 19.8% | 196 | 11.6% |
| Had trauma team | | 650 | 9.8% | 508 | 54.8% | 77 | 30.0% | 68 | 4.0% |
| Had CT scan | | 5669 | 85.2% | 743 | 80.2% | 246 | 95.7% | 1080 | 63.9% |
| Had surgery | | 234 | 3.5% | 403 | 43.5% | 26 | 10.1% | 207 | 12.2% |
| Had interventional radiology | | 7 | 0.1% | 12 | 1.3% | 0 | 0.0% | 1 | 0.1% |
| Intubated and ventilated | | 511 | 8.5% | 73 | 14.7% | 39 | 18.1% | 16 | 1.2% |
| Admitted to critical care | | 231 | 3.8% | 267 | 42.4% | 26 | 11.4% | 75 | 5.4% |
| Patient underwent transfer | | 6653 | 100.0% | 927 | 100.0% | 257 | 100.0% | 1691 | 100.0% |
| Rehabilitation prescription | | 481 | 14.6% | 730 | 92.3% | 67 | 82.7% | 115 | 12.0% |
| Survived to 30 days | | 6601 | 99.2% | 925 | 99.8% | 257 | 100.0% | 1684 | 99.6% |
| Length of hospital stay (days) | | 1 | 1 - 9 | 15 | 8 - 25 | 1 | 1 - 5 | 2 | 1 - 9 |
| Length of critical care stay (days) | | 1 | 1 - 4 | 8 | 3 - 19 | 8 | 4 - 13 | 3 | 1 - 6 |
| Discharged to own home | | 167 | 71.7% | 22 | 42.3% | 6 | 85.7% | 32 | 54.2% |

| Supplementary table 10: Discharge destination by trauma centre status | | | | | | | | |
|---|---|---|---|---|---|---|---|---|
| | **Trauma Unit** 92159 (72.4%) | | **Major Trauma Centre** 35175 (27.6%) | | **Comparison** | | | |
| | n | % | n | % | Diff | OR | 95% CI | Sig |
| Own home | 52635 | 57.1% | 21012 | 59.7% | 2.6% | 1.114 | 1.08 -1.142 | <0.000 |
| Nursing home | 9439 | 10.2% | 2808 | 8.0% | 2.2% | 0.76 | 0.728-0.795 | <0.000 |
| Rehabilitation | 8727 | 9.5% | 4606 | 13.1% | 3.6% | 1.44 | 1.387-1.496 | <0.000 |
| Other acute hospital | 8368 | 9.1% | 1408 | 4.0% | 5.1% | 0.418 | 0.394-0.442 | <0.000 |
| Mortuary | 7006 | 7.6% | 3787 | 10.8% | 3.2% | 1.466 | 1.407-1.529 | <0.000 |
| Home (of relative or other) | 2636 | 2.9% | 504 | 1.4% | 1.5% | 0.049 | 0.448-0.543 | <0.000 |
| Other institution | 2493 | 2.7% | 889 | 2.5% | 0.2% | 0.933 | 0.863-1.008 | 0.076 |
| Missing destination | 292 | 0.3% | 41 | 0.1% | 0.2% | 0.367 | 0.265-0.509 | <0.000 |
| Social care | 239 | 0.3% | 58 | 0.2% | 0.1% | 0.635 | 0.477-0.847 | 0.001 |
| Alive in hospital at 30 days | 201 | 0.2% | 3 | 0.0% | 0.2% | 0.039 | 0.012-0.122 | <0.000 |
| No fixed abode | 123 | 0.1% | 59 | 0.2% | 0.1% | 1.257 | 0.921-1.715 | 0.172 |
| **Dichotomised discharge destination** | | | | | | | | |
| Own home | 52635 | 57.1% | 21012 | 59.7% | 2.6% | 1.06 | 1.029-1.091 | <0.000 |
| Formal or informal care | 23534 | 25.5% | 8865 | 25.2% | 0.3% | 0.943 | 0.917-0.972 | <0.000 |
| Excluded* | 15990 | 17.4% | 5298 | 15.1% | | | - | |

Significance values are Chi-squared with Yates' continuity correction.
**Diff**: difference. **OR**: odds ratio. **CI**: confidence intervals. **Sig**: significance.
*Died, remained in hospital at 30 days (censored), transferred, discharged to no fixed abode, missing destination.

| Supplementary Table 11: Logistic regression model for discharge destination. | | | | | | |
|---|---|---|---|---|---|---|
| | | B | Sig. | Exp(B) | 95% CI (lower) | 95% CI (upper) |
| Age | 16 - 44 (reference) | | | | | |
| | 45 - 54 | 1.42 | 0.04 | 4.14 | 1.06 | 16.19 |
| | 55 - 64 | 2.34 | 0.00 | 10.41 | 3.98 | 27.27 |
| | 65 - 74 | 3.36 | 0.00 | 28.87 | 10.49 | 79.43 |
| | 75 - 85 | 3.74 | 0.00 | 42.14 | 22.14 | 80.21 |
| | >85 | 1.76 | 0.00 | 5.79 | 3.15 | 10.65 |
| Sex | Female (reference) | | | | | |
| | Male | 0.09 | 0.00 | 1.10 | 1.06 | 1.13 |
| Age * Sex interaction | 16 - 44 * Male (reference) | | | | | |
| | 45 - 54 * Male | -0.03 | 0.05 | 0.97 | 0.95 | 1.00 |
| | 55 - 64 * Male | -0.04 | 0.00 | 0.96 | 0.94 | 0.97 |
| | 65 - 74 * Male | -0.06 | 0.00 | 0.94 | 0.93 | 0.96 |
| | 75 - 85 * Male | -0.06 | 0.00 | 0.94 | 0.93 | 0.95 |
| | >85 * Male | -0.04 | 0.00 | 0.96 | 0.95 | 0.97 |
| MCCI | 0 (reference) | | | | | |
| | 1 - 5 | -0.42 | 0.00 | 0.66 | 0.63 | 0.68 |
| | 6 - 10 | -0.54 | 0.00 | 0.59 | 0.56 | 0.61 |
| | >10 | -0.69 | 0.00 | 0.50 | 0.47 | 0.54 |
| Anticoagulated | Not anticoagulated (ref) | | | | | |
| | Prescribed anticoagulant | 0.03 | 0.23 | 1.03 | 0.98 | 1.07 |
| ISS | (ISS/10)2 | -0.17 | 0.00 | 0.84 | 0.81 | 0.87 |
| | Log(ISS/10) x (ISS/10)2 | 0.07 | 0.00 | 1.07 | 1.04 | 1.10 |
| GCS | 15 (reference) | | | | | |
| | 3 | -1.54 | 0.00 | 0.22 | 0.17 | 0.28 |
| | 4 - 5 | -1.53 | 0.00 | 0.22 | 0.14 | 0.34 |
| | 6 - 8 | -1.30 | 0.00 | 0.27 | 0.22 | 0.34 |
| | 9 - 12 | -0.99 | 0.00 | 0.37 | 0.34 | 0.41 |
| | 13 - 14 | -0.58 | 0.00 | 0.56 | 0.54 | 0.59 |
| Shock | No shock (reference) | | | | | |
| | Shock | -0.10 | 0.00 | 0.90 | 0.87 | 0.94 |
| Injury distribution | Abdomen (reference) | | | | | |
| | Chest | -0.27 | 0.04 | 0.76 | 0.59 | 0.99 |
| | Face | -0.07 | 0.71 | 0.94 | 0.67 | 1.31 |
| | Head | -0.25 | 0.06 | 0.78 | 0.60 | 1.01 |
| | Limbs | -1.32 | 0.00 | 0.27 | 0.21 | 0.35 |
| | Multiple | -1.05 | 0.00 | 0.35 | 0.27 | 0.45 |
| | Other | -1.09 | 0.00 | 0.34 | 0.18 | 0.64 |
| | Spine | -0.67 | 0.00 | 0.51 | 0.40 | 0.66 |
| Trauma centre status | Trauma Unit (reference) | | | | | |
| | Major Trauma Centre | 0.06 | 0.00 | 1.07 | 1.03 | 1.10 |
| Constant | | 3.52 | 0.00 | 33.64 | 25.66 | 44.12 |
| Model performance and characteristics: Cox & Snell $R^2$ 0.131, Nagelkerke $R^2$ 0.186, AUROC 0.744 (95% CI 0.739 - 0.749, p <0.000) | | | | | | |

| Supplementary Table 12: Cox Regression model for length of stay | | | | | | |
|---|---|---|---|---|---|---|
| Variable | Categories | Coefficient | Significance | Exp(B) | 95% CI (lower) | 95% CI (upper) |
| Trauma centre status | Trauma Unit (reference) | | | | | |
| | Major Trauma Centre | -0.09 | 0.00 | 0.91 | 0.90 | 0.93 |
| Age | 16 - 44 (reference) | | | | | |
| | 45 - 54 | -0.19 | 0.00 | 0.83 | 0.78 | 0.87 |
| | 55 - 64 | -0.29 | 0.00 | 0.75 | 0.72 | 0.79 |
| | 65 - 74 | -0.59 | 0.00 | 0.55 | 0.53 | 0.58 |
| | 75 - 85 | -0.75 | 0.00 | 0.47 | 0.45 | 0.50 |
| | >85 | -0.82 | 0.00 | 0.44 | 0.42 | 0.46 |
| Sex | Female (reference) | | | | | |
| | Male | 0.00 | 0.92 | 1.00 | 0.94 | 1.05 |
| MCCI | 0 (reference) | | | | | |
| | 1 - 5 | -0.24 | 0.00 | 0.78 | 0.77 | 0.80 |
| | 6 - 10 | -0.32 | 0.00 | 0.72 | 0.71 | 0.74 |
| | >10 | -0.44 | 0.00 | 0.64 | 0.62 | 0.66 |
| Anticoagulated | Not anticoagulated (ref) | | | | | |
| | Prescribed anticoagulant | 0.00 | 0.66 | 1.00 | 0.99 | 1.02 |
| GCS | 15 (reference) | | | | | |
| | 3 | -0.97 | 0.00 | 0.38 | 0.34 | 0.43 |
| | 4 - 5 | -0.89 | 0.00 | 0.41 | 0.34 | 0.49 |
| | 6 - 8 | -0.74 | 0.00 | 0.48 | 0.43 | 0.53 |
| | 9 - 12 | -0.49 | 0.00 | 0.62 | 0.59 | 0.65 |
| | 13 - 14 | -0.18 | 0.00 | 0.84 | 0.82 | 0.85 |
| Shock | No shock (reference) | | | | | |
| | Shock | -0.10 | 0.00 | 0.90 | 0.89 | 0.92 |
| Injury distribution | Abdomen (reference) | | | | | |
| | Chest | 0.06 | 0.14 | 1.06 | 0.98 | 1.16 |
| | Face | 0.34 | 0.00 | 1.40 | 1.25 | 1.56 |
| | Head | 0.19 | 0.00 | 1.21 | 1.12 | 1.32 |
| | Limbs | -0.37 | 0.00 | 0.69 | 0.63 | 0.75 |
| | Multiple | -0.30 | 0.00 | 0.74 | 0.68 | 0.81 |
| | Other | -0.11 | 0.42 | 0.89 | 0.68 | 1.18 |
| | Spine | -0.29 | 0.00 | 0.75 | 0.69 | 0.81 |
| ISS | $(ISS/10)^2$ | -0.16 | 0.00 | 0.85 | 0.84 | 0.86 |
| | $Log(ISS/10) \times (ISS/10)^2$ | 0.07 | 0.00 | 1.07 | 1.06 | 1.08 |
| Age * Sex interaction | 16 - 44 * Male (reference) | | | | | |
| | 45 - 54 * Male | -0.09 | 0.01 | 0.91 | 0.85 | 0.98 |
| | 55 - 64 * Male | -0.13 | 0.00 | 0.88 | 0.82 | 0.93 |
| | 65 - 74 * Male | -0.04 | 0.25 | 0.96 | 0.90 | 1.03 |
| | 75 - 85 * Male | -0.02 | 0.49 | 0.98 | 0.92 | 1.04 |
| | >85 * Male | -0.01 | 0.78 | 0.99 | 0.93 | 1.05 |

**Exp(B)**: Odds ratio. **CI**: confidence interval.
